# Causal Association of Inflammatory Bowel Disease on Anorexia Nervosa: A Two-sample Mendelian Randomization Study

**DOI:** 10.1101/2022.08.21.22279045

**Authors:** Rui Jiang, Ruijie Zeng, Zewei Zhuo, Huihuan Wu, Qi Yang, Jingwei Li, Felix W Leung, Weihong Sha, Hao Chen

## Abstract

**Background:** Anorexia nervosa (AN) is a severe and complex psychiatric disorder with the highest mortality rate of all psychiatric disorders. Previous observational studies have comprehensively indicated an association between inflammatory bowel disease (IBD) and AN, but the causal effect remains unclear. We aimed to test the hypothesis that a causal relationship exists between IBD and the risk of AN.

**Methods:** Based on published open genome-wide association study summary statistics, the single nucleotide polymorphisms (SNPs) of IBD, including ulcerative colitis (UC) and Crohn’s disease (CD), were selected as genetic instrumental variables. Two-sample Mendelian randomization (MR) analyses were conducted to infer the causal relationship between IBD and AN. The methods included inverse variance weighted (IVW), MR-Egger regression, weighted median estimator, weighted mode and MR Pleiotropy RESidual Sum and Outlier (MR-PERSSO). Furthermore, various sensitivity analyses were conducted to assess the robustness of our results.

**Results:** The primary analysis IVW suggested that genetically predicted IBD were positively casually associated with AN risk (N_SNP_ = 12, odds ratio [OR]: 1.143, 95% confidence interval [CI]: 1.032-1.266, *P* = 0.01). In subgroups, consistent positive causal associations with AN risk might show in UC (N_SNP_ = 6, OR: 1.153, 95% CI: 1.022-1.301, *P* = 0.021), and CD (N_SNP_ = 4, OR: 1.153, 95% CI: 1.008-1.319, *P* = 0.037). NO heterogeneity and pleiotropy were presented through sensitivity analysis and guaranteed credible and robust estimates.

**Conclusion:** Our study identifies that patients diagnosed with IBD, including UC and CD, could be causally associated with an increased risk of AN. It may have clinical benefits for early detection of AN and clinical decisions on management of IBD patients.

## 1. Introduction

Anorexia nervosa (AN) is a severe and complex psychiatric disorder with the highest mortality rate of all psychiatric disorders[1]. Patients with AN restrict caloric intake due to a strong fear of weight gain, resulting in significantly low body weight and severe malnutrition. Prolonged and severe malnutrition can cause widespread organ dysfunction, disability or even death[2], severely impairing the patient’s physical and psychological functioning. In a recent large US cohort study, the lifetime prevalence of AN was 1.42% in adult women and a lower relative prevalence of 0.12% in adult men. Genetic, neurobiological, developmental and environmental factors all contribute toward the pathogenesis of AN[3]. Previous studies have revealed that AN is strongly associated with gastrointestinal changes and symptoms, and conversely, gastrointestinal symptoms may also influence the development of AN[4]. Inflammatory bowel disease (IBD), mainly comprising ulcerative colitis (UC) and Crohn’s disease (CD), is a multifactorial and relapsing gastrointestinal inflammatory disease. Clinically, the presentations of IBD and AN overlap: reduced BMI, female predominance, similar age of onset, contributing to diagnostic confusion and delay in treatment[5]. AN with comorbid IBD increased certain difficulty of precise diagnosis and might lead to the failure of correct and effective treatment, resulting in long-term unalleviated condition and worse prognosis [6, 7].

Epidemiological studies have suggested interesting but inconclusive clinical correlations between IBD and AN. Compared with healthy controls, patients with IBD showed a higher prevalence of eating disorder symptoms including AN[8]. Strong links between CD (odds ratio [OR]:1.97 (1.42-2.67), *P* <0.001), UC (OR: 2.25 (1.57-3.12), *P* <0.001) and AN were shown in a UK record linkage cohort study of eating disorders and autoimmunity diseases[9]. An increased risk of immune-mediated gastrointestinal disease (including UC and CD) was found in AN patients compared with control subjects[10]. A previous diagnosis of AN had a risk ratio of 1.44 (1.05, 1.97) for any IBD, 1.60 (1.04, 2.46) for CD, and 1.66 (1.15, 2.39) for UC, but an IBD diagnosis was not significantly associated AN[11].

Traditional observational studies could not establish a causal relationship between IBD and AN, because of limited sample size, chance of reverse causality, confounding and measurement error[12]. To avoid these shortcomings and verify our hypothesis, we propose the use of Mendelian randomization (MR) method.MR is an approach to deduce estimates for the causal effect of risk factors on disease outcomes by utilizing genetic variants, primarily single nucleotide polymorphisms (SNPs), as instrumental variables (IVs)[13]. Using Genome-wide association study (GWAS) summary data, two-sample MR method was conducted on the association of IBD and AN with IVs. In addition, we implemented the same two-sample MR analysis in two-subtypes of IBD, UC and CD.

## 2. Methods

### 2.1 Data Resources

Summary-level genetic instruments for IBD (3,753 cases and 210,300 controls), CD (2,701 cases and 215,806 controls), and UC (2,056 cases and 210,300 controls) were obtained from the FinnGen database, a collection of genomic and health data from approximately 500,000 Finnish biobank participants[14].

A large-scale GWAS meta-analysis of AN involved 14,477 individuals incorporating 3,495 subjects with AN and 10,982 healthy controls[15]. Derived from the Eating Disorders Work Group of the Psychiatric Genomics Consortium (PGC-ED), all participants recruited to our study were European ancestry. Details of the GWAS datasets were briefly listed in Supplementary Table S1. All summary statistics were publicly available, and no additional ethical approval or informed consent for this study was required.

### 2.2 Selection of Genetic Instrumental Variables

SNPs selected in this research were statistically significant (P < 5.0×10-8) and independent (R2 < 0.001) at the genetic level. Strict selection criteria (r2= 0.001, kb=10000) were adopted to maximally reduce the interference of linkage disequilibrium (LD). Effect allele mismatch may bias causal effect estimates in the opposite direction[16]. For harmonization, the orientation of the alleles was corrected and the palindromic SNPs were excluded to ensure that genetic associations were expressed correctly with the same allele on both strands. Furthermore, the F-statistics calculated as (beta/se)^2^ was computed to evaluate the strength of IVs, and values > 10 represented strong instruments. We confirmed that the exposures and outcomes studies applied in our two-sample MR analyses did not involve overlapping participants by reading the literature of data sources.

### 2.3 Mendelian Randomization Analysis

We conducted two-sample MR analyses with inverse variance weighted (IVW) as the main method to appraise the potential causality of associations between IBD and AN. In addition to IVW, complementary analyses including MR-Egger regression, weighted median estimator, weighted mode and MR Pleiotropy RESidual Sum and Outlier (MR-PERSSO)[17] were utilized to provide more credible evidence for the association. Wald ratio calculated MR effect estimates for each SNP. Meta-analysis of Wald ratios was performed using IVW. The weighted median method can provide reliable and more efficient estimates when over 50% of the weight derived from valid IVs. Weighted mode-based method indicated an unbiased estimate with the largest weights associated with the valid IVs[18]; MR Pleiotropy RESidual Sum and Outlier (MR-PRESSO) performs an IVW MR for detection and correction for heterogeneity via the global test and outlier test respectively. All results are expressed as odds ratio including 95% confidence interval. Scatter plots were also generated to visualize the individual SNP and combined estimates from each MR method.

### 2.4 Sensitivity Analysis

To provide a more credible estimate, sensitivity analyses were performed to assess the heterogeneity and pleiotropy of our study as well as potential genetic outliers. The Cochran’s Q test was calculated to examine heterogeneity across individual SNPs in the IVW and MR egger methods. *P* < 0.05 was considered to be significantly heterogeneous. As for horizontal pleiotropy, MR-Egger intercept estimates and MR-PRESSO outlier analysis were applied to test and correct possible horizontal pleiotropic outliers in the analysis. Funnel plots were also produced for visual examination of possible directional pleiotropy by observing symmetry. The leave-one-out analyses excluded each SNP from the analysis in turn, and performed a two-sample MR analysis using the remaining SNPs as IVs to verify whether the overall causal effect is driven by an individual SNP. Based on the principle of MR, we considered the association as causal when (1) at least the main results of IVW were significant; (2) insignificant heterogeneity or a random effects model was employed in case of heterogeneity; (3) the estimates adjusted for pleiotropy suggested null effects; (4) results were not completely dominated by any individual SNP.

### 2.5 Statistical analysis

All analyses were undertaken by R software (version 4.1.1) using the R package “TwoSampleMR”. A two-tailed *P* < 0.05 means the difference is statistically significant in both MR analyses and sensitivity estimates.

## 3. Results

### 3.1 Genetic Instrumental variants

After removing linkage disequilibrium and palindromes, 2 SNPs were excluded and 16 SNPs for IBD were for the subsequent analysis, whereas 5 and 8 genetic instruments were eventually retained for CD and UC. The corresponding F-statistic of each adopted SNP is > 10 (range from 31.10 to 50.31), indicating that the causality results we obtained can ignore the possible influence of weak instrumental variables. Detailed information on stringently selected genetic instruments for subsequent MR analysis was listed in Supplementary Table S2.

### 3.2 Causal Effect of IBD on AN

Primary IVW estimates suggested that patients with IBD typically have an increasing incidence of AN (N_SNP_ = 12, odds ratio (OR): 1.143, 95% CI: 1.032-1.266, *P* = 0.01; Figure 2). In subgroups, consistent positive causal association with AN risk showed in UC (N_SNP_ = 6, OR: 1.153, 95% CI: 1.022-1.301, *P* = 0.021), and CD (N_SNP_ = 4, OR: 1.153, 95% CI: 1.008-1.319, *P* = 0.037). Scatter plots (Figure 3) showed that the effect estimation obtained by the four MR methods all pointed to the same direction, and indicate a positive correlation. The individual SNP final selected for these causal results were displayed in forest plots (Supplementary Figure S1).

**Figure 1.**
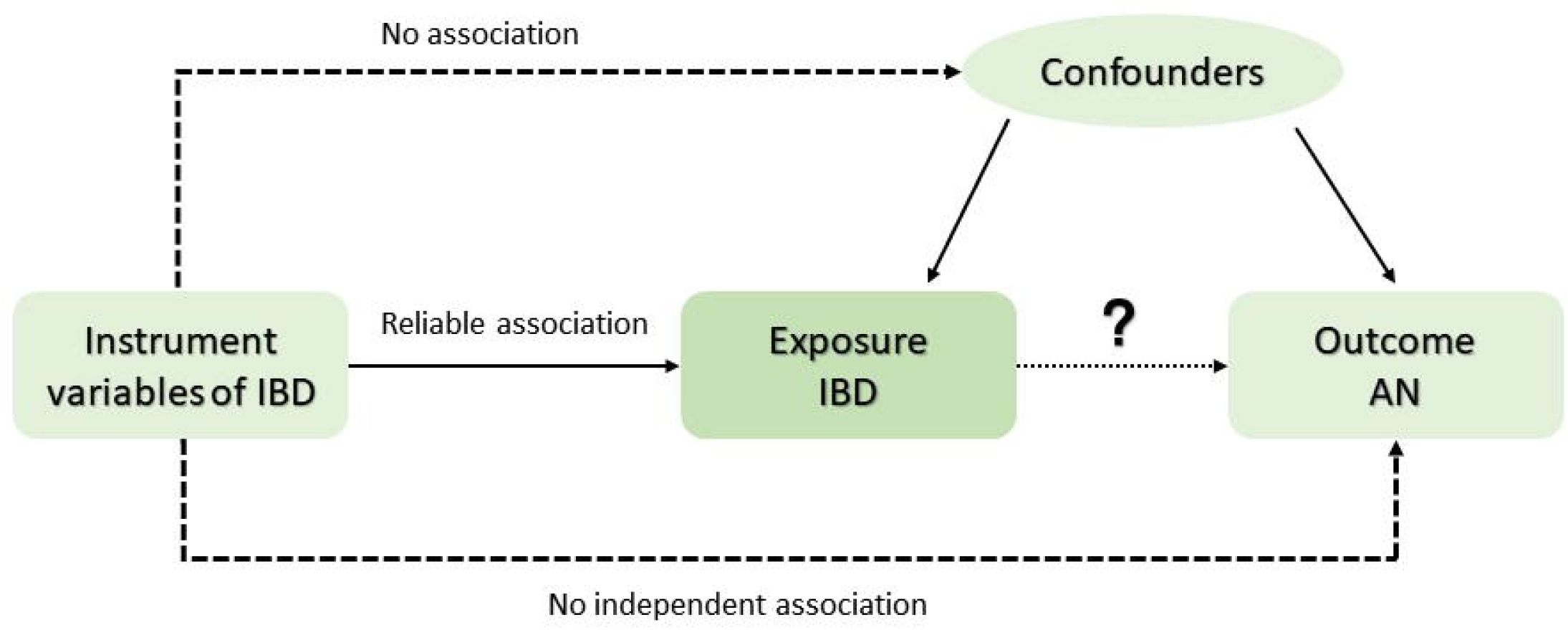
Three rigorous assumptions underpin the principle and validity of MR analysis. The genetic instruments should be: 1) associated with the IBD; 2) affected AN only through the IBD; and 3) independent of any confounders in the relation between IBD and AN.

**Figure 2.**
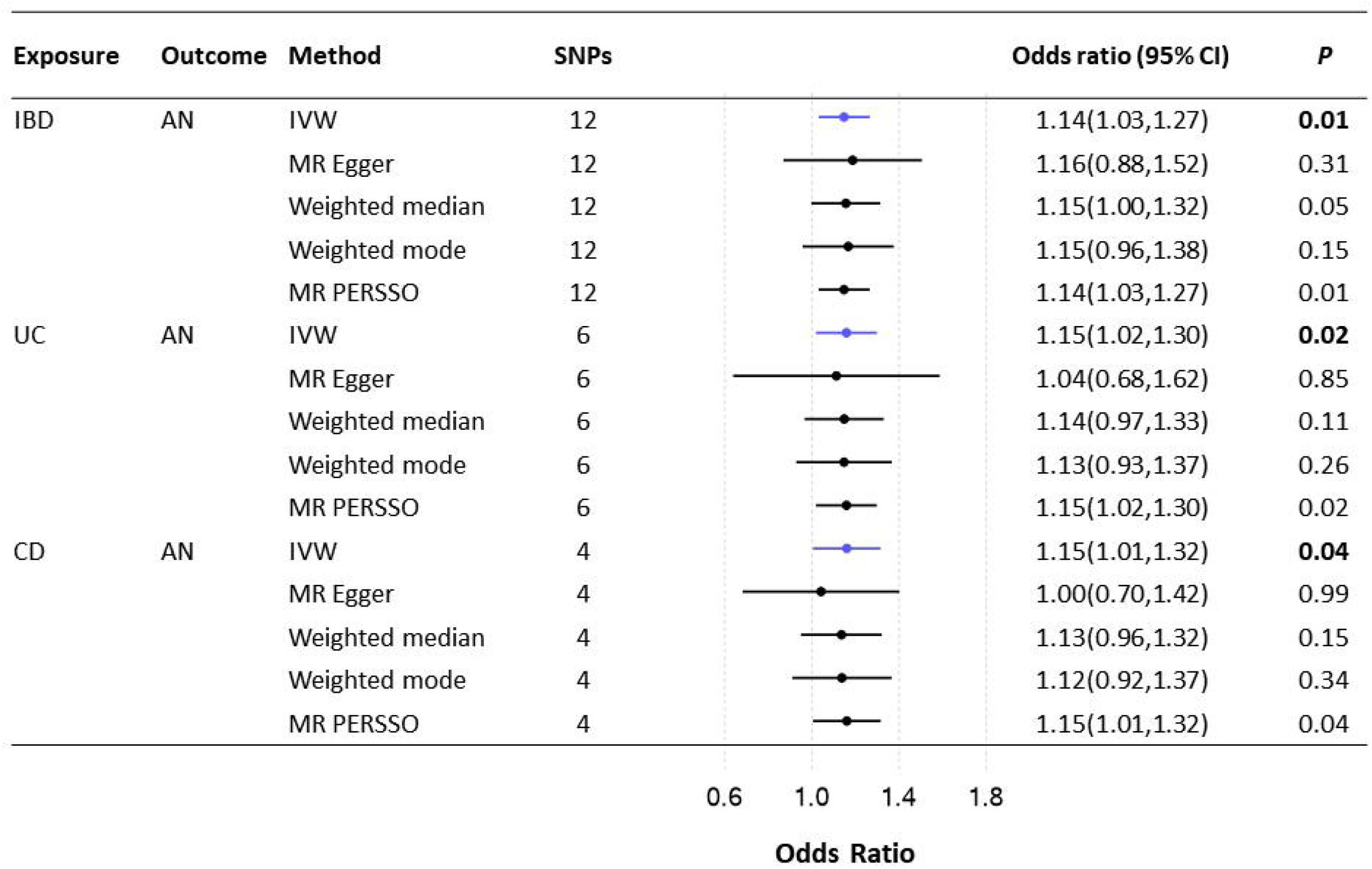
MR analysis results for the causal associations of IBD on AN.

**Figure 3.**
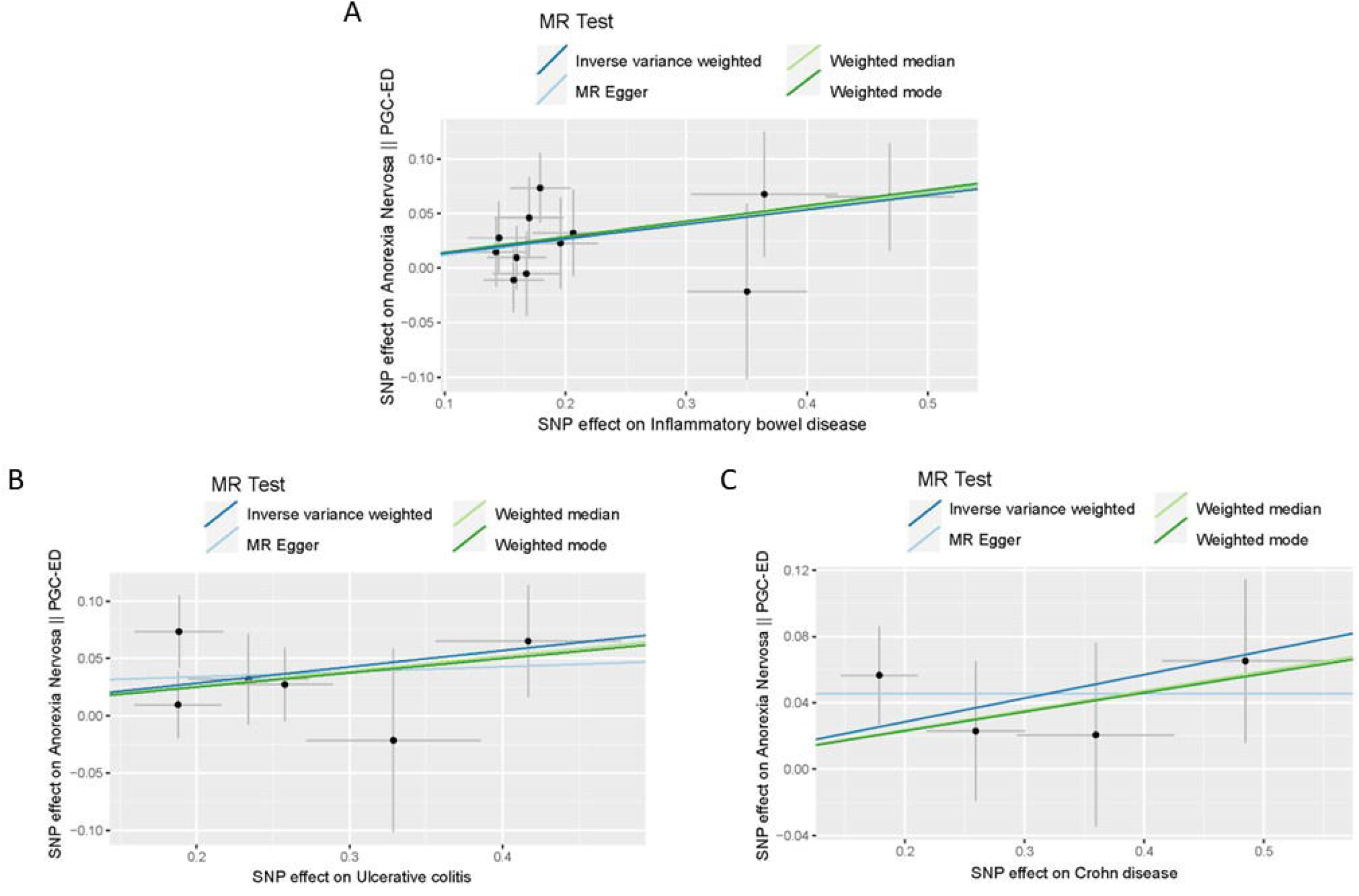
Scatter plots of causal effect between IBD, including UC and CD, on risk of AN. A. IBD on AN; B. UC on AN; C. CD on AN.

### 3.3 Sensitivity analysis

The corresponding *F*-statistic (range from 31.10-50.31) of IVs for IBD suggested that strong instruments were selected in our MR analysis (Supplementary Table S2). Cochran’s Q test of IVW (*P*_IBD_= 0.90; *P*_UC_ = 0.65; *P*_CD_ =0.67; Supplementary Table S3) indicated no-existence of heterogeneity, the results remained similar with those estimates from MR-PRESSO Global test (*P*_IBD_= 0.90; *P*_UC_ = 0.38; *P*_CD_ =0.75; Supplementary Table S3). No obvious outlier SNP was identified in the MR-PRESSO Outlier test for IBD, UC and CD on AN. In the MR-Egger regression analysis, no directional pleiotropy effect was observed (IBD: *P*_intercept = 0.92; UC: *P*_intercept = 0.67; CD: *P*_intercept = 0.48; Supplementary Table S3), which indicated that the estimate from the IVW analysis is not biased. Funnel plots showed that the points representing the causal association effect of each SNP are symmetrically distributed, indicating that the association is less affected by potential bias (Supplementary Figure S2). In the leave-one-out analyses, no potential driving SNP was observed in estimated causal effect between IBD and AN when we systematically dropped the individual SNP and repeated the MR analyses in turn (Supplementary Figure S3). However, the leave-one-out plots for UC and CD on AN did not present robust results. The aforementioned sensitivity analysis indicated generally robustness and consistency to the causal associations.

## 4. Discussion

Overall, our results render relatively strong evidence to support that, compared with healthy individuals, it is possible that having IBD could contribute to the occurrence and development of AN. And the same positive causal association might be found for UC and CD on risk of AN. It may have clinical benefits for early detection of AN and clinical decisions on management of IBD patients.

In the past research, Hedman et al. observed a statistically significant association between gastrointestinal-related CD and AN in women based on the Swedish population[19]. In 2018, Collins et al reported that a young woman diagnosed with CD also experienced relapse and continued weight loss with prompt treatment, and subsequently developed AN[20]. Another case report described a patient with long-standing CD and AN who had significant improvement in pre-existing AN symptom after receiving anti-TNF-a therapy corresponding to IBD[21]. Our study aimed to further explore the causal association between IBD and AN based on a series of studies on the epidemiological findings.

Reflecting a high suicide rate, AN remains a serious problem in modern medicine in terms of prediction, diagnosis, and treatment due to unclear etiology and complex clinical manifestations, including various physical and psychiatric symptoms. Both IBD[22] and AN[23] are relapsing and multifactorial diseases in which genetics, immunity and gut microbiota play important roles. Several studies have suggested that changes in dietary patterns and gastrointestinal symptoms in IBD patients may be predisposing factors for AN[5, 24]. IBD positively reinforces eating disorder behaviors in patients with comorbid anorexia nervosa, contributing to the worsening of the eating disorder and providing barriers to its treatment. Likewise, psychopathology examinations revealed significantly elevated levels of anxiety, depression, and eating disorder psychopathology in the AN patients compared with healthy controls[25]. Intestinal inflammation alters brain function, including effects on induced mood disorders and appetite. In addition, significant gut microbial dysbiosis was both observed in patients with IBD and AN[26, 27]. Dysregulation of the gut microbiota affects the body’s energy balance and neurohormone production, possibly affecting the central nervous system through many interactions along the gut-brain axis[28, 29]. Molecularly, it has been suggested that pro-inflammatory cytokines have an effect on anorexic properties by signaling the regulation of appetite[21, 30]. Due to the paucity of pathogenesis studies, it is certainly difficult to clear the definitive molecular mechanism that IBD is a direct cause or effect of observed AN symptom, and further studies were required. The definitely causal relationship prompts clinical practitioners to remain vigilant and suspicious of the clinical manifestations of AN in patients with IBD. Frequent screening of mental and dietary status of IBD patients to reduce the occurrence of AN or early detection and treatment.

The strength of the present study is that two-sample MR has stronger evidence of causality than observational epidemiology, and is less prone to biases from confounding and reverse causation. Rigorous criteria were utilized in the selection of IVs to ensure that MR assumptions were satisfied. Moreover, we also conducted a series of sensitivity analyses, and did not find any heterogeneity and multiple effects affecting the results between IBD and AN, which effectively avoided the interference of false negatives, and underpin the accuracy of our results. Also, some limitations and deficiency existed in our study. First, MR analysis was restricted primarily to populations of European ancestry, and our findings may not be generalizable in all ethnicities. Second, the largest GWAS summary data of AN were utilized in our study, but the quantity of SNPs was still insufficient. Limited number of SNPs possibly explaining the inconsistency of leave-one-out plots for UC and CD on AN in our study. Further prospective studies with larger sample sizes are needed to provide more adequate statistical power to validate the causal association. Third, observational epidemiology studies had shown a higher prevalence of AN and IBD comorbidities in young women. However, the published summary data we used do not have detailed in age and sex stratification. A more precise stratification analysis of age and gender in disease requires larger sample sizes and detailed GWAS data. Admittedly, though we demonstrated a causal association between IBD and AN, additional studies were still needed to explore the potential role and molecular mechanisms underlying the observed associations.

## 5 Conclusion

In summary, our two-sample MR method results confirm the hypothesis that patients with IBD are more prone to develop the risk of AN. It is suggested that special attention should be paid to the clinical manifestations related to AN when patients are diagnosed with IBD, and timely preventive measures should be taken for patients with related symptoms to reduce the risk of anorexia and improve the well-being of patients. In addition, subsequent biological mechanisms and experimental studies are expected to provide a reasonable interpretation for our findings and verify the potential molecular mechanisms for the effect of IBD on the development of AN.

## Data Availability

All data produced in the present work are contained in the manuscript

## Acknowledgment

We thank all patients donated their tissue samples and the FinnGen and PGC-ED Consortium generously make their data openly available.

## Conflict of Interest

The authors have declared no conflicts of interest.

## Funding

This work was funded by the National Natural Science Foundation of China (82171698, 82170561, 81300279, 81741067), the Natural Science Foundation for Distinguished Young Scholars of Guangdong Province (2021B1515020003), Natural Science Foundation of Guangdong Province (2022A1515012081), the Climbing Program of Introduced Talents and High-level Hospital Construction Project of Guangdong Provincial People’s Hospital (DFJH201803, KJ012019099, KJ012021143, KY012021183), the Foreign Distinguished Teacher Program of Guangdong Science and Technology Department (KD0120220129).

## Author Contributions

Felix W Leung, Weihong Sha and Hao Chen designed and supervised the study; Rui Jiang, Ruijie Zeng and Huihuan Wu carried out the statistical analyses; Zewei Zhuo, Qi Yang, Jingwei Li conducted the study; Rui Jiang, Ruijie Zeng contributed to wrote the manuscript; All authors had read and approved the final version of manuscript.

## Data Availability

All GWAS summary data involved in this study are publicly available data. IBD, UC and CD data are available at https://www.finngen.fi/fi AN data is available at https://gwas.mrcieu.ac.uk/datasets/ieu-a-1186/

## Figure Legends

Supplementary Figure S1. The individual SNP final selected for these causal results were displayed in forest plots. A. IBD on AN; B. UC on AN; C. CD on AN.

Supplementary Figure S2. Funnel plots for individual causal effect estimate. A. IBD on AN; B. UC on AN; C. CD on AN.

Supplementary Figure S3. Leave-one-out analysis plots. A. IBD on AN; B. UC on AN; C. CD on AN.

Supplementary Table S1. Sources of the GWAS datasets utilized in our study.

Supplementary Table S2. Detailed SNPs stringently selected genetic instruments for subsequent MR analysis.

Supplementary Table S3. Sensitivity analyses results of MR-Egger intercept test, Cochran’s Q test and MR-PRESSO.

## Notes

### Competing Interest Statement

The authors have declared no competing interest.

## References

1 Arcelus J, Mitchell AJ, Wales J, Nielsen S. Mortality rates in patients with anorexia nervosa and other eating disorders. A meta-analysis of 36 studies. Arch Gen Psychiatry. 2011, 68:724–731 [PMID: 21727255] [doi:10.1001/archgenpsychiatry.2011.74]

2 Zipfel S, Giel KE, Bulik CM, Hay P, Schmidt U. Anorexia nervosa: aetiology, assessment, and treatment. Lancet Psychiatry. 2015, 2:1099–1111 [PMID: 26514083] [doi:10.1016/S2215-0366(15)00356-9]

3 Force USPST, Davidson KW, Barry MJ, Mangione CM, Cabana M, Chelmow D, Coker TR, Davis EM, Donahue KE, Jaen CR, Kubik M, Li L, Ogedegbe G, Pbert L, Ruiz JM, Silverstein M, Stevermer J, Wong JB. Screening for Eating Disorders in Adolescents and Adults: US Preventive Services Task Force Recommendation Statement. JAMA. 2022, 327:1061–1067 [PMID: 35289876] [doi:10.1001/jama.2022.1806]

4 Santonicola A, Gagliardi M, Guarino MPL, Siniscalchi M, Ciacci C, Iovino P. Eating Disorders and Gastrointestinal Diseases. Nutrients. 2019, 11 [PMID: 31842421] [PMC6950592] [doi:10.3390/nu11123038]

5 Ilzarbe L, Fabrega M, Quintero R, Bastidas A, Pintor L, Garcia-Campayo J, Gomollon F, Ilzarbe D. Inflammatory Bowel Disease and Eating Disorders: A systematized review of comorbidity. J Psychosom Res. 2017, 102:47–53 [PMID: 28992897] [doi:10.1016/j.jpsychores.2017.09.006]

6 Erdur L, Kallenbach-Dermutz B, Lehmann V, Zimmermann-Viehoff F, Kopp W, Weber C, Deter HC. Somatic comorbidity in anorexia nervosa: First results of a 21-year follow-up study on female inpatients. Biopsychosoc Med. 2012, 6:4 [PMID: 22300749] [PMC3299644] [doi:10.1186/1751-0759-6-4]

7 Bayle FJ, Bouvard MP. Anorexia nervosa and Crohn’s disease dual diagnosis: a case study. Eur Psychiatry. 2003, 18:421–422 [PMID: 14680721] [doi:10.1016/j.eurpsy.2003.01.002]

8 Satherley R, Howard R, Higgs S. Disordered eating practices in gastrointestinal disorders. Appetite. 2015, 84:240–250 [PMID: 25312748] [doi:10.1016/j.appet.2014.10.006]

9 Wotton CJ, James A, Goldacre MJ. Coexistence of eating disorders and autoimmune diseases: Record linkage cohort study, UK. Int J Eat Disord. 2016, 49:663–672 [PMID: 27333941] [doi:10.1002/eat.22544]

10 Raevuori A, Haukka J, Vaarala O, Suvisaari JM, Gissler M, Grainger M, Linna MS, Suokas JT. The increased risk for autoimmune diseases in patients with eating disorders. PLoS One. 2014, 9:e104845 [PMID: 25147950] [PMC4141740] [doi:10.1371/journal.pone.0104845]

11 Larsen JT, Yilmaz Z, Vilhjálmsson BJ, Thornton LM, Benros ME, Musliner KL, Consortium EDWGotPG, Werge T, Hougaard DM, Mortensen PB, Bulik CM, Petersen LV. Anorexia nervosa and inflammatory bowel diseases—Diagnostic and genetic associations. JCPP Advances. 2021, 1:e12036 [PMID: [doi:https://doi.org/10.1002/jcv2.12036]

12 Davey Smith G, Hemani G. Mendelian randomization: genetic anchors for causal inference in epidemiological studies. Hum Mol Genet. 2014, 23:R89–98 [PMID: 25064373] [PMC4170722] [doi:10.1093/hmg/ddu328]

13 Emdin CA, Khera AV, Kathiresan S. Mendelian Randomization. JAMA. 2017, 318:1925–1926 [PMID: 29164242] [doi:10.1001/jama.2017.17219]

14 Kurki MI, Karjalainen J, Palta P, Sipilä TP, Kristiansson K, Donner K, Reeve MP, Laivuori H, Aavikko M, Kaunisto MA, Loukola A, Lahtela E, Mattsson H, Laiho P, Della Briotta Parolo P, Lehisto A, Kanai M, Mars N, Rämö J, Kiiskinen T, Heyne HO, Veerapen K, Rüeger S, Lemmelä S, Zhou W, Ruotsalainen S, Pärn K, Hiekkalinna T, Koskelainen S, Paajanen T, Llorens V, Gracia-Tabuenca J, Siirtola H, Reis K, Elnahas AG, Aalto-Setälä K, Alasoo K, Arvas M, Auro K, Biswas S, Bizaki-Vallaskangas A, Carpen O, Chen C-Y, Dada OA, Ding Z, Ehm MG, Eklund K, Färkkilä M, Finucane H, Ganna A, Ghazal A, Graham RR, Green E, Hakanen A, Hautalahti M, Hedman Å, Hiltunen M, Hinttala R, Hovatta I, Hu X, Huertas-Vazquez A, Huilaja L, Hunkapiller J, Jacob H, Jensen J-N, Joensuu H, John S, Julkunen V, Jung M, Junttila J, Kaarniranta K, Kähönen M, Kajanne RM, Kallio L, Kälviäinen R, Kaprio J, Kerimov N, Kettunen J, Kilpeläinen E, Kilpi T, Klinger K, Kosma V-M, Kuopio T, Kurra V, Laisk T, Laukkanen J, Lawless N, Liu A, Longerich S, Mägi R, Mäkelä J, Mäkitie A, Malarstig A, Mannermaa A, Maranville J, Matakidou A, Meretoja T, Mozaffari SV, Niemi ME, Niemi M, Niiranen T, O’Donnell CJ, Obeidat Me, Okafo G, Ollila HM, Palomäki A, Palotie T, Partanen J, Paul DS, Pelkonen M, Pendergrass RK, Petrovski S, Pitkäranta A, Platt A, Pulford D, Punkka E, Pussinen P, Raghavan N, Rahimov F, Rajpal D, Renaud NA, Riley-Gillis B, Rodosthenous R, Saarentaus E, Salminen A, Salminen E, Salomaa V, Schleutker J, Serpi R, Shen H-y, Siegel R, Silander K, Siltanen S, Soini S, Soininen H, Sul JH, Tachmazidou I, Tasanen K, Tienari P, Toppila-Salmi S, Tukiainen T, Tuomi T, Turunen JA, Ulirsch JC, Vaura F, Virolainen P, Waring J, Waterworth D, Yang R, Nelis M, Reigo A, Metspalu A, Milani L, Esko T, Fox C, Havulinna AS, Perola M, Ripatti S, Jalanko A, Laitinen T, Mäkelä T, Plenge R, McCarthy M, Runz H, Daly MJ, Palotie A. FinnGen: Unique genetic insights from combining isolated population and national health register data. medRxiv. 2022:2022.2003.2003.22271360 [PMID: [doi:10.1101/2022.03.03.22271360]

15 Duncan L, Yilmaz Z, Gaspar H, Walters R, Goldstein J, Anttila V, Bulik-Sullivan B, Ripke S, Eating Disorders Working Group of the Psychiatric Genomics C, Thornton L, Hinney A, Daly M, Sullivan PF, Zeggini E, Breen G, Bulik CM. Significant Locus and Metabolic Genetic Correlations Revealed in Genome-Wide Association Study of Anorexia Nervosa. Am J Psychiatry. 2017, 174:850–858 [PMID: 28494655] [PMC5581217] [doi:10.1176/appi.ajp.2017.16121402]

16 Hartwig FP, Davies NM, Hemani G, Davey Smith G. Two-sample Mendelian randomization: avoiding the downsides of a powerful, widely applicable but potentially fallible technique. Int J Epidemiol. 2016, 45:1717–1726 [PMID: 28338968] [PMC5722032] [doi:10.1093/ije/dyx028]

17 Ong JS, MacGregor S. Implementing MR-PRESSO and GCTA-GSMR for pleiotropy assessment in Mendelian randomization studies from a practitioner’s perspective. Genet Epidemiol. 2019, 43:609–616 [PMID: 31045282] [PMC6767464] [doi:10.1002/gepi.22207]

18 Hartwig FP, Davey Smith G, Bowden J. Robust inference in summary data Mendelian randomization via the zero modal pleiotropy assumption. Int J Epidemiol. 2017, 46:1985–1998 [PMID: 29040600] [PMC5837715] [doi:10.1093/ije/dyx102]

19 Hedman A, Breithaupt L, Hubel C, Thornton LM, Tillander A, Norring C, Birgegard A, Larsson H, Ludvigsson JF, Savendahl L, Almqvist C, Bulik CM. Bidirectional relationship between eating disorders and autoimmune diseases. J Child Psychol Psychiatry. 2019, 60:803–812 [PMID: 30178543] [doi:10.1111/jcpp.12958]

20 Collins A, Nolan E, Hurley M, D’Alton A, Hussey S. Anorexia Nervosa Complicating Pediatric Crohn Disease-Case Report and Literature Review. Front Pediatr. 2018, 6:283 [PMID: 30356737] [PMC6189420] [doi:10.3389/fped.2018.00283]

21 Solmi M, Santonastaso P, Caccaro R, Favaro A. A case of anorexia nervosa with comorbid Crohn’s disease: beneficial effects of anti-TNF-alpha therapy? Int J Eat Disord. 2013, 46:639-641 [PMID: 23813727] [doi:10.1002/eat.22153]

22 Basso PJ, Fonseca MT, Bonfa G, Alves VB, Sales-Campos H, Nardini V, Cardoso CR. Association among genetic predisposition, gut microbiota, and host immune response in the etiopathogenesis of inflammatory bowel disease. Braz J Med Biol Res. 2014, 47:727–737 [PMID: 25075576] [PMC4143199] [doi:10.1590/1414-431x20143932]

23 Gorwood P, Blanchet-Collet C, Chartrel N, Duclos J, Dechelotte P, Hanachi M, Fetissov S, Godart N, Melchior JC, Ramoz N, Rovere-Jovene C, Tolle V, Viltart O, Epelbaum J. New Insights in Anorexia Nervosa. Front Neurosci. 2016, 10:256 [PMID: 27445651] [PMC4925664] [doi:10.3389/fnins.2016.00256]

24 Mascolo M, Geer B, Feuerstein J, Mehler PS. Gastrointestinal comorbidities which complicate the treatment of anorexia nervosa. Eat Disord. 2017, 25:122–133 [PMID: 27869566] [doi:10.1080/10640266.2016.1255108]

25 Borgo F, Riva A, Benetti A, Casiraghi MC, Bertelli S, Garbossa S, Anselmetti S, Scarone S, Pontiroli AE, Morace G, Borghi E. Microbiota in anorexia nervosa: The triangle between bacterial species, metabolites and psychological tests. PLoS One. 2017, 12:e0179739 [PMID: 28636668] [PMC5479564] [doi:10.1371/journal.pone.0179739]

26 Nikolova VL, Hall MRB, Hall LJ, Cleare AJ, Stone JM, Young AH. Perturbations in Gut Microbiota Composition in Psychiatric Disorders: A Review and Meta-analysis. JAMA Psychiatry. 2021, 78:1343–1354 [PMID: 34524405] [PMC8444066] [doi:10.1001/jamapsychiatry.2021.2573]

27 Seitz J, Belheouane M, Schulz N, Dempfle A, Baines JF, Herpertz-Dahlmann B. The Impact of Starvation on the Microbiome and Gut-Brain Interaction in Anorexia Nervosa. Front Endocrinol (Lausanne). 2019, 10:41 [PMID: 30809191] [PMC6379250] [doi:10.3389/fendo.2019.00041]

28 Karakula-Juchnowicz H, Pankowicz H, Juchnowicz D, Valverde Piedra JL, Malecka-Massalska T. Intestinal microbiota - a key to understanding the pathophysiology of anorexia nervosa? Psychiatr Pol. 2017, 51:859–870 [PMID: 29289966] [doi:10.12740/PP/65308]

29 Roubalova R, Prochazkova P, Papezova H, Smitka K, Bilej M, Tlaskalova-Hogenova H. Anorexia nervosa: Gut microbiota-immune-brain interactions. Clin Nutr. 2020, 39:676–684 [PMID: 30952533] [doi:10.1016/j.clnu.2019.03.023]

30 Collins SM. Interrogating the Gut-Brain Axis in the Context of Inflammatory Bowel Disease: A Translational Approach. Inflamm Bowel Dis. 2020, 26:493–501 [PMID: 31970390] [PMC7054772] [doi:10.1093/ibd/izaa004]

