## supplemental Files for "Causal Association of Inflammatory Bowel Disease on Anorexia Nervosa: A Two-sample Mendelian Randomization Study"

### Supplementary Figure S1

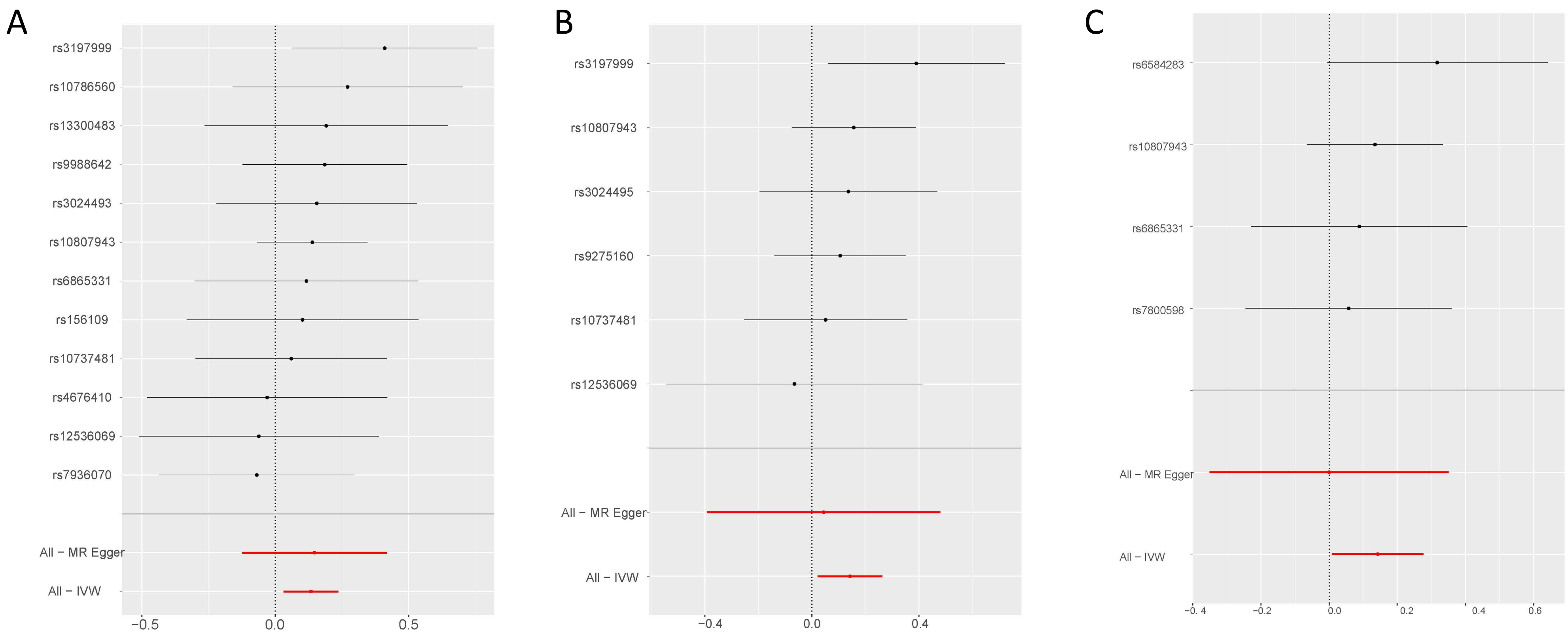

Supplementary Figure S2

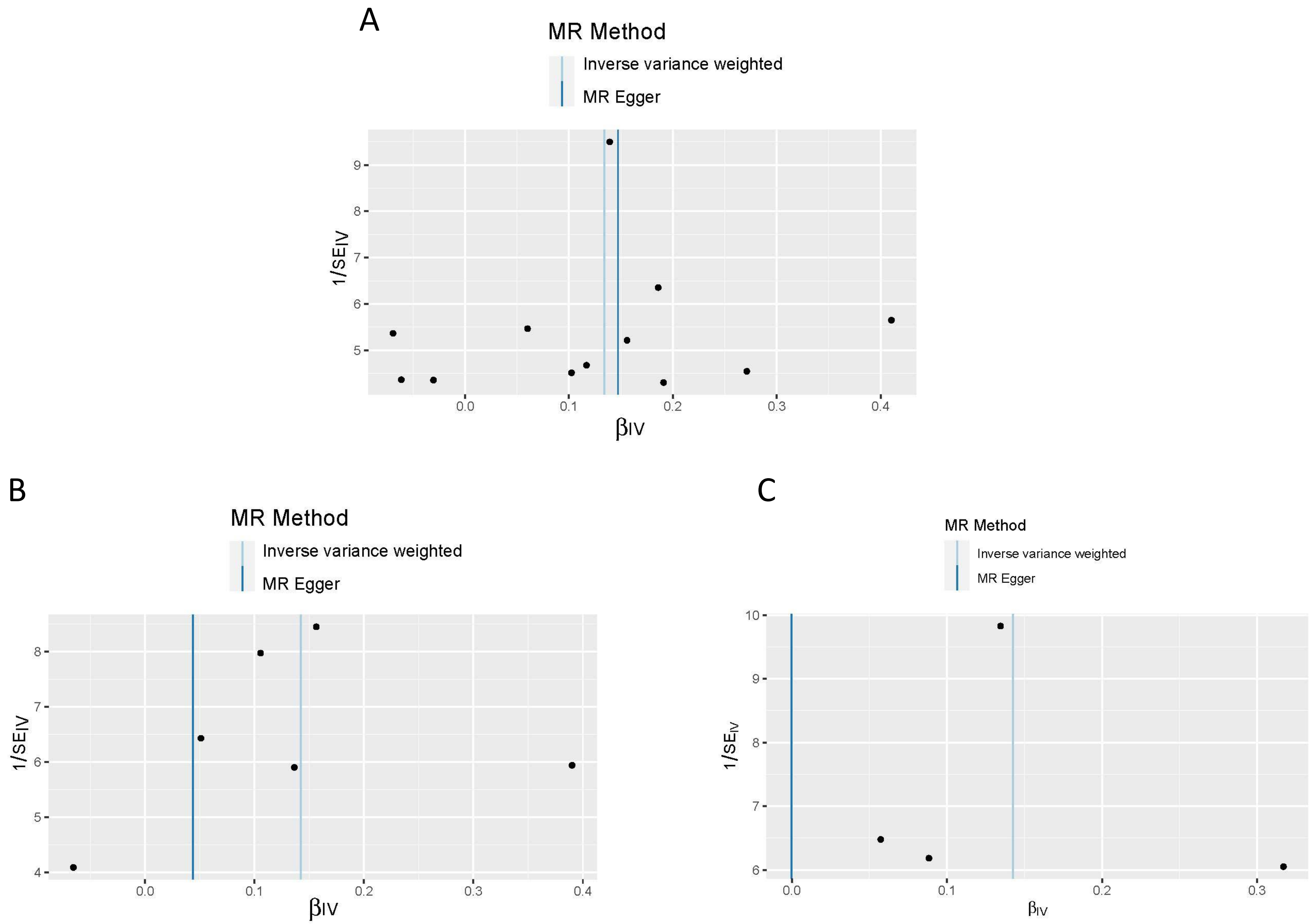

### Supplementary Figure S3

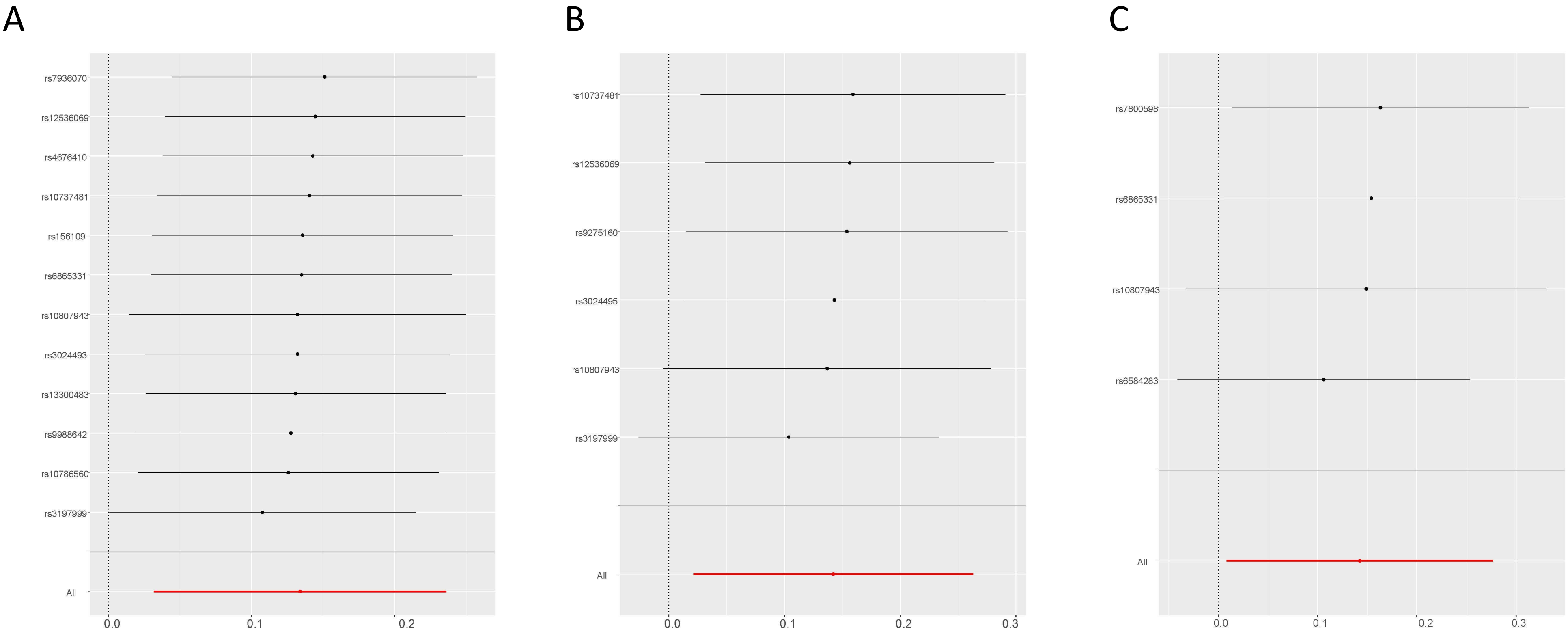

**Supplementary Table S1. Sources of the GWAS datasets utilized in our study.**

| Traits | Significant level | Data sources | Sample size (cases/controls) | Ancestry | Reference |
|---|---|---|---|---|---|
| IBD | 5.00E-08 | FinnGen | 3,753/210,300 | European | Mitja I. Kurki et,al |
| CD | 5.00E-08 | FinnGen | 2,056/210,300 | European | Mitja I. Kurki et,al |
| UC | 5.00E-08 | FinnGen | 2,701/215,806 | European | Mitja I. Kurki et,al |
| AN | 5.00E-08 | PGC-ED | 3,495/10,982 | European | Duncan L et,al |

IBD: inflammatory bowel disease; CD: Crohn's disease; UC: ulcerative colitis; AN: anorexia nervosa; PGC-ED: Eating Disorders Work Group of the Psychiatric Genomics Consortium; GWAS： Genome-wide association study.

**Supplementary Table S2. Detailed SNPs stringently selected genetic instruments for subsequent MR analysis.**

| SNP | EA/ non-EA | Chr | Location | Beta (SE) | F statistics | *P* value | AN [Beta (SE)] |
|---|---|---|---|---|---|---|---|
| **IBD** | | | | | | | |
| rs10737481 | G/T | 1 | 20171514 | 0.16(0.02) | 42.78 | 5.90E-11 | 0.06(0.18) |
| rs10786560 | A/G | 10 | 101315166 | 0.17(0.03) | 35.09 | 3.10E-09 | 0.27(0.22) |
| rs10807943 | C/T | 7 | 5340664 | -0.47(0.05) | 78.96 | 6.83E-19 | 0.14(0.11) |
| rs12536069 | C/T | 7 | 4909470 | 0.35(0.05) | 50.31 | 1.36E-12 | -0.06(0.23) |
| rs13300483 | T/C | 9 | 117643362 | 0.15(0.03) | 31.10 | 2.55E-08 | 0.19(0.23) |
| rs156109 | T/C | 5 | 131626611 | 0.14(0.03) | 31.77 | 1.78E-08 | 0.10(0.22) |
| rs3024493 | A/C | 1 | 206943968 | 0.21(0.03) | 37.33 | 1.03E-09 | 0.16(0.19) |
| rs3197999 | A/G | 3 | 49721532 | 0.18(0.03) | 50.91 | 8.85E-13 | 0.41(0.18) |
| rs4676410 | A/G | 2 | 241563739 | 0.17(0.03) | 36.92 | 1.23E-09 | -0.03(0.23) |
| rs6865331 | G/A | 5 | 40326099 | 0.20(0.03) | 40.19 | 2.38E-10 | 0.12(0.21) |
| rs7936070 | T/G | 11 | 76293527 | 0.16(0.02) | 40.51 | 1.93E-10 | -0.07(0.19) |
| rs9988642 | C/T | 1 | 67726104 | -0.36(0.06) | 36.22 | 1.75E-09 | 0.19(0.16) |
| **UC** | | | | | | | |
| rs10737481 | G/T | 1 | 20171514 | 0.19(0.03) | 43.73 | 3.53E-11 | 0.05(0.16) |
| rs10807943 | C/T | 7 | 5340664 | -0.42(0.06) | 46.97 | 7.13E-12 | 0.16(0.12) |
| rs12536069 | C/T | 7 | 4909470 | 0.33(0.06) | 33.00 | 9.01E-09 | -0.07(0.24) |
| rs3024495 | T/C | 1 | 206942413 | 0.23(0.04) | 35.39 | 2.70E-09 | 0.14(0.17) |
| rs3197999 | A/G | 3 | 49721532 | 0.19(0.03) | 41.92 | 9.73E-11 | 0.39(0.17) |
| rs9275160 | A/G | 6 | 32652620 | -0.26(0.03) | 66.88 | 2.61E-16 | 0.11(0.13) |
| **CD** | | | | | | | |
| rs10807943 | C/T | 7 | 5340664 | -0.48(0.07) | 48.6 | 3.16E-12 | -0.07(0.05) |
| rs6584283 | C/T | 10 | 101290301 | -0.18(0.03) | 30.54 | 3.38E-08 | -0.06(0.03) |
| rs6865331 | G/A | 5 | 40326099 | 0.26(0.04) | 39.94 | 2.67E-10 | 0.02(0.04) |
| rs7800598 | A/G | 7 | 4862677 | 0.36(0.07) | 29.85 | 4.77E-08 | 0.02(0.06) |

IBD: inflammatory bowel disease; CD: Crohn's disease; UC: ulcerative colitis; AN: anorexia nervosa; SNP: single nucleotide polymorphism; EA: effect allele; Chr: chromosome; SE: standard error; MR: Mendelian randomization.

**Supplementary Table S3. Sensitivity analyses results of MR-Egger intercept test, Cochran's Q test and MR-PRESSO.**

| Exposure | IVW | | Weighted Median | MR-Egger | | MR PRESSO | | |
|---|---|---|---|---|---|---|---|---|
| | OR (95% CI) | Q (*p value*) | OR (95% CI) | OR (95% CI) | Intercept (*p value*) | outlier | global test RSSobs | *P* |
| IBD | 1.14(1.03,1.27) | 5.63(0.90) | 1.15(1.00,1.32) | 1.16(0.88,1.52) | -0.003(0.92) | 0 | 6.59 | 0.90 |
| UC | 1.15(1.02,1.30) | 3.34(0.65) | 1.14(0.97,1.33) | 1.04(0.68,1.62) | 0.03(0.67) | 0 | 6.07 | 0.38 |
| CD | 1.15(1.01,1.32) | 1.54(0.67) | 1.13(0.96,1.32) | 1.00(0.70,1.42) | 0.05(0.48) | 0 | 2.28 | 0.75 |

IBD: inflammatory bowel disease; CD: Crohn's disease; UC: ulcerative colitis; AN: anorexia nervosa; CI: confidence interval; OR: odds ratio; IVW: inverse variance weighted; MR PRESSO: Mendelian Randomization Pleiotropy RESidual Sum and Outlier.
